# Multisectoral prioritization of zoonotic diseases in India: A One Health perspective

**DOI:** 10.1101/2024.02.26.24303393

**Authors:** Simmi Tiwari, Indranil Roy, Monal Daptardar, Ruchi Singh, Anshuman Mishra, Richa Kedia, Harmesh Manocha, Mayank Diwedi, Amlesh Dwivedi, Gaurish Shukla, Ajit Shewale, Tushar Nale, Dipti Mishra, R.P. Sharma, Daniel Garcia, Runa H. Gokhale, Meghna Desai, Sujeet Singh

## Abstract

**Introduction:** To tackle the risk of emerging and re-emerging diseases, it is critical for countries with limited resources to prioritize endemic and emerging zoonotic diseases of greatest national concern. One Health is an integrated, unifying approach that aims to sustainably balance and optimize the health of people, animals, and ecosystems. In India, as a first step towards a multi-disciplinary, multi-sectoral, One Health approach to preventing and detecting zoonotic disease outbreaks, a national-level multi-stakeholder zoonotic disease prioritization workshop was organized to identify a list of zoonotic diseases of greatest national concern for India.

**Methods:** We followed the Good Reporting of a Mixed Methods Study (GRAMMS) guidelines to finalize a list of priority zoonotic diseases through a participatory action research approach involving 50 experts in zoonotic diseases. We used a prioritization process based on the U.S. Centers for Disease Control and Prevention’s semi-quantitative One Health Zoonotic Disease Prioritization (OHZDP) Process, with modifications per country need.

**Results:** We ranked forty zoonotic diseases based on five criteria: severity of illness in humans, the economic burden of the diseases, pandemic potential, capacity for prevention and control, and potential for introduction or increased transmission in India. The final list of zoonotic diseases ranked in the order of national significance includes the following top ten priority zoonotic diseases: Zoonotic Influenza (Zoonotic Influenza A viruses), Anthrax, Japanese Encephalitis, Leptospirosis, Brucellosis, Dengue, Rabies, Scrub typhus, Plague, and Crimean-Congo hemorrhagic fever. We conducted a sensitivity analysis to assess the impact of each criterion on the prioritized list; this analysis showed minimal changes in ranking for the top ten diseases.

**Conclusion:** For the successful adoption of One Health practices in India, multi-sectoral collaboration is critical at all levels – national, state, and provincial. This collaborative prioritization process conducted at the national level has the potential to catalyse such efforts and enhance zoonotic disease prevention and detection efforts at the state and local levels across India.

## Introduction

India is one of 12 mega biodiverse countries in the world, with 11% of the world’s flora in about 2.4% of its land mass [1]. Intensification of agriculture, rapid urbanization, and population growth have altered ecosystems such that the natural balance is towards increased animal-human association [2], which brings increased risk to wildlife and humans from emerging and re-emerging infectious diseases [3,4]. As such, India has been identified as a hotspot for the transmission of both known and novel infectious agents between animals and people [5].

One Health is an integrated, unifying approach that aims to sustainably balance and optimize the health of people, animals, and ecosystems. It recognizes that the health of humans, domestic and wild animals, plants, and the wider environment (including ecosystems) are closely linked and inter-dependent [6]. Zoonoses (infections that are transmitted between animals and humans) currently in circulation globally include human immunodeficiency virus-1 (HIV-1), Ebola virus, highly pathogenic avian influenza (HPAI) viruses, and the novel coronaviruses - severe acute respiratory syndrome (SARS) coronavirus, Middle East respiratory syndrome (MERS) coronavirus, and severe acute respiratory syndrome coronavirus 2 (SARS-CoV-2). An estimated 20% of all human illnesses and deaths in less developed countries are attributable to endemic zoonoses [7]. The COVID-19 pandemic, driven by SARS-CoV-2 and its several variants, is a recent example of a zoonotic threat. The COVID-19 pandemic caused an unprecedented level of morbidity and mortality across the globe and fulfilled the prediction of global health experts that another pandemic with the speed and severity of the 1918 influenza epidemic was a matter *“not of if, but of when”* [8]. Addressing such a One Health challenge requires an understanding of risks that exist at the interface between humans, animals, and their environments [9].

A robust understanding of the pathogen ecology of natural host and human–host interactions is required in order to inform surveillance and public health interventions for preventing and/or mitigating future zoonotic spillover events [10]. Prioritization is required in order to ensure the strengthening of public health systems and efficient use of existing resources through a collaborative, multisectoral, transdisciplinary One Health approach [11]. Zoonotic disease prioritization enables countries to plan and calibrate their activities through a coordinated effort across the human, animal, wildlife, and environmental health sectors. Zoonotic disease prioritization involves the use of various tools incorporating qualitative, semi-quantitative, and quantitative methods [12–19]. The U.S. Centers for Disease Control and Prevention’s (US-CDC) One Health Zoonotic Disease Prioritization (OHZDP) Process is a tool used to identify priority zoonotic diseases and develop next steps and action plans to tackle them [20].

In India, zoonotic infectious diseases such as cholera (*Vibrio cholerae* serogroup O139), sylvatic plague, Nipah virus disease, diphtheria, Chandipura virus disease, Chikungunya virus infection, HPAI A (H5N1), novel influenza A (H1N1), Crimean-Congo haemorrhagic fever (CCHF), leptospirosis, anthrax, Kala-azar, scrub typhus, acute encephalitis syndrome and Kyasanur forest disease (KFD), are reported through the Integrated Disease Surveillance Program (IDSP). IDSP is a national health programme started in 2009 designed to strengthen and maintain a decentralized laboratory-based disease surveillance system for epidemic prone diseases, to monitor disease trends, and to detect and respond to outbreaks [21]. Disease outbreaks reported from India include rabies (ongoing), Chandipura virus (*Rhabdoviridae)* (1965, 2003, 2004, 2007) [22,23], and Chikungunya virus disease (1963, 1973) [24].

A modified OHZDP Process was used in India in 2019 for the prioritization of zoonotic disease specific to the Indian city of Ahmedabad [25], however similar prioritization has not been done at the national level. As a first step towards a national One Health approach to the prevention and detection of zoonotic diseases, a national-level multi-stakeholder, multi-sectoral zoonotic disease prioritization workshop was organized in Jaipur, in the state of Rajasthan, by the provincial health department of Rajasthan and the National Centre for Disease Control (NCDC), Government of India, with the support of the US-CDC, India. The objectives of the workshop were to identify a list of zoonotic diseases of greatest national concern for India and to develop cohesive, sustainable strategies for the prevention and control of these priority zoonoses.

## Materials and methods

This study to finalize a list of priority zoonotic diseases for India, through a participatory action research approach, followed the Good Reporting of a Mixed Methods Study (GRAMMS) guidelines [26]. A mixed methods approach was necessary to incorporate both the qualitative and quantitative components adopted before, during, and after the three-day national, multi-stakeholder workshop (Fig 1). Discussions involved 50 experts in zoonotic diseases from institutes including the Ministry of Fisheries, Animal Husbandry & Dairying, the Ministry of Agriculture and Farmers Welfare, the Ministry of Environment, Forest and Climate Change, the Ministry of Health and Family Welfare, state governments, private agencies, and national and international organizations

**Fig 1.**
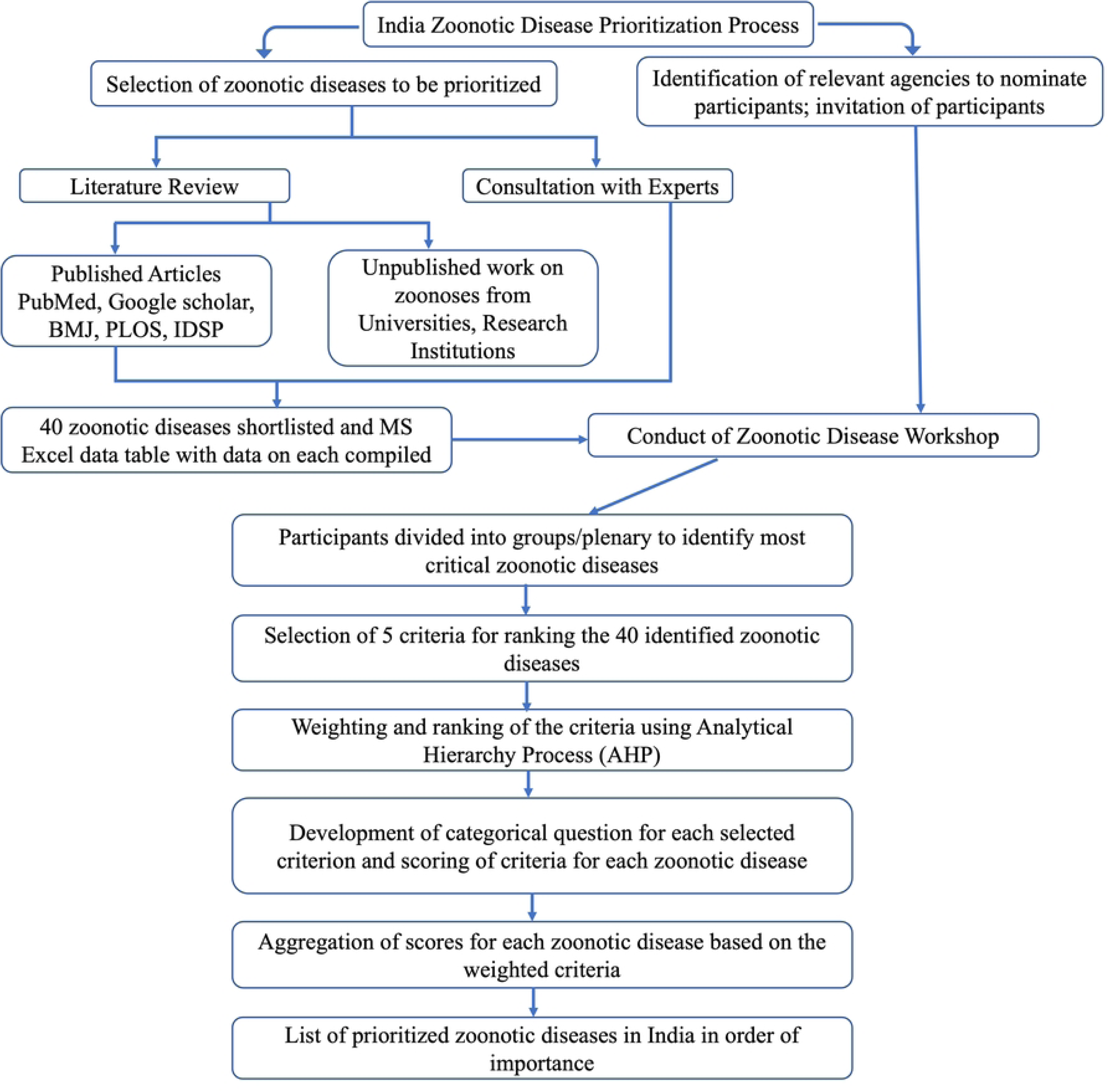
Flow of Workshop.

US-CDC’s One Health Zoonotic Disease Prioritization (OHZDP) Process [20] was modified for country-specific requirements and used for this prioritization. The modified prioritization process was completed using the following six-step sequence of methods:

### Step 1: Selection of experts from the field and potential priority zoonotic diseases

Experts were identified by first determining stakeholder organizations. A stakeholder was any organization, institution, network, or group, involved in activities pertaining to zoonotic diseases at the national or international level (Fig 2). Initially, a formal communication was sent to the State Health Departments, Department of Animal Husbandry (Ministry of Animal Husbandry, Dairying, and Fisheries), Department of Wildlife (Ministry of Environment and Forests), State Medical Colleges, Indian Council of Agriculture Research (ICAR, Ministry of Agriculture and Farmers Welfare), Indian Council of Medical Research (ICMR, Ministry of Health and Family Welfare), veterinary universities, and international agencies in India, to nominate or identify experts from these sectors at the provincial and federal level. Experts from these agencies were identified for participation in the workshop and were categorised as voters (N=33), advisors (N=10), and facilitators (N=7). The roles and responsibilities of each of these categories were as below:

**Fig 2.**
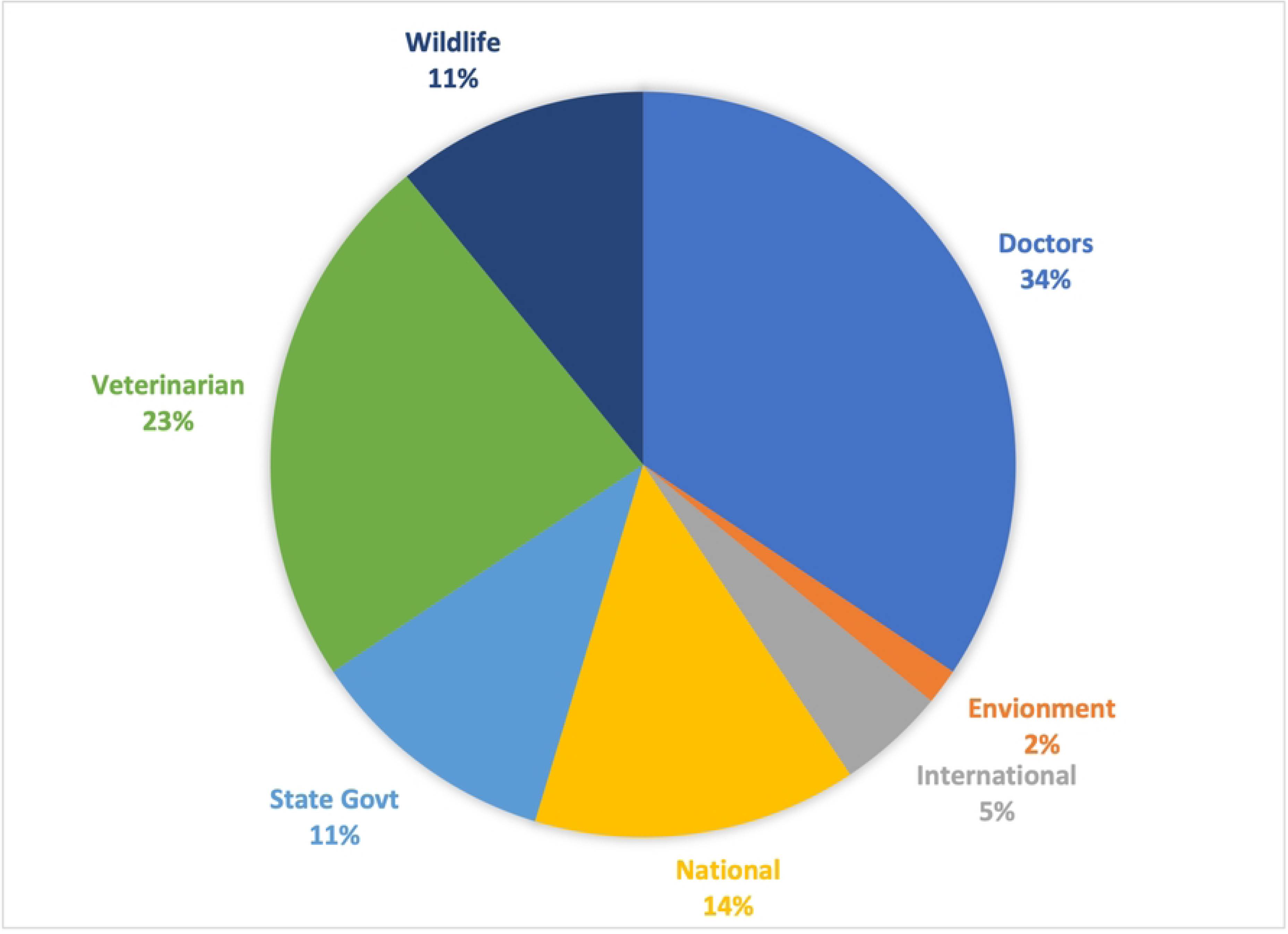
Experts in the zoonotic disease prioritization workshop.

#### Voting members

Voting members were subject matter experts from government public health, veterinary, wildlife, environment, and food sectors, as well as from international and private agencies. They were individuals with technical knowledge and programmatic experience in the field of zoonotic diseases. Their role during the workshop was to provide key inputs as per the selected criteria and to support the prioritization of zoonotic diseases.

#### Advisors

Advisors were senior administrators and technical experts working in the public health, veterinary, wildlife, environment, and food sectors. They provided expertise during discussions with voting members to develop a multisectoral One Health zoonotic disease prevention, detection, and response plan for India.

#### Facilitators

Facilitators were experts with advanced computer skills and an understanding of the various methods used for prioritization, including the analytic hierarchy process (AHP). They steered the decision making of voting members through objective, evidence-based data gathered from the literature review. The AHP is a multi-criteria decision making (MCDM) method that is flexible and easy to use. It is used to identify and/or prioritize the net benefit of health interventions [27], in this case the prioritization of zoonotic diseases according to their overall impact on human life.

Three rounds of preworkshop online discussion were held to acquaint the voting members, advisors, and facilitators with the steps, process, and expected outcomes of the workshop. During these discussions, a list of 40 potential priority zoonoses was developed through a focused literature review and discussions Table 2. The literature review included reports available under Government health programs and peer-reviewed publications indexed on PubMed with publication dates including January 2000 through January 2020 (using search terms “zoonotic disease” OR “zoonoses”, OR “infectious disease” AND “India”).

### Step 2: Development of criteria for ranking the zoonotic diseases

Prior to the workshop the advisors, facilitators and most of the voting members identified in Step 1 debated the criteria to be included, the supporting questions and responses, and the scores, until consensus was achieved. Through this process five criteria, along with categorical questions that would enable weighting and ranking for prioritization of zoonotic diseases, were decided upon. Prioritization documents produced by other countries were used as a reference. The discussion was led by NCDC. The five criteria that were developed were: the severity of disease in humans; the economic burden of disease; pandemic or epidemic potential; potential of transmission; and capacity for prevention and control in humans and the animal health sector. These criteria were presented at the workshop for discussion and ultimately used during the prioritization process Table 3.

### Step 3: National multi-stakeholder zoonotic disease prioritization workshop

A three-day national multi-stakeholder zoonotic disease prioritization workshop was held in Jaipur, Rajasthan, from 10-12 February 2020. This workshop included the 50 facilitators, advisors, and voting members described above, all experts in the fields of epidemiology, microbiology, parasitology, public health, veterinary public health, environmental health, and international health Table **1**. The day one agenda included the finalization of the weightage and ranking of criteria, the day two agenda included the finalization of zoonotic disease rankings, and the day three agenda included a workshop summary and action plan (Fig 1).

**Table 1.**
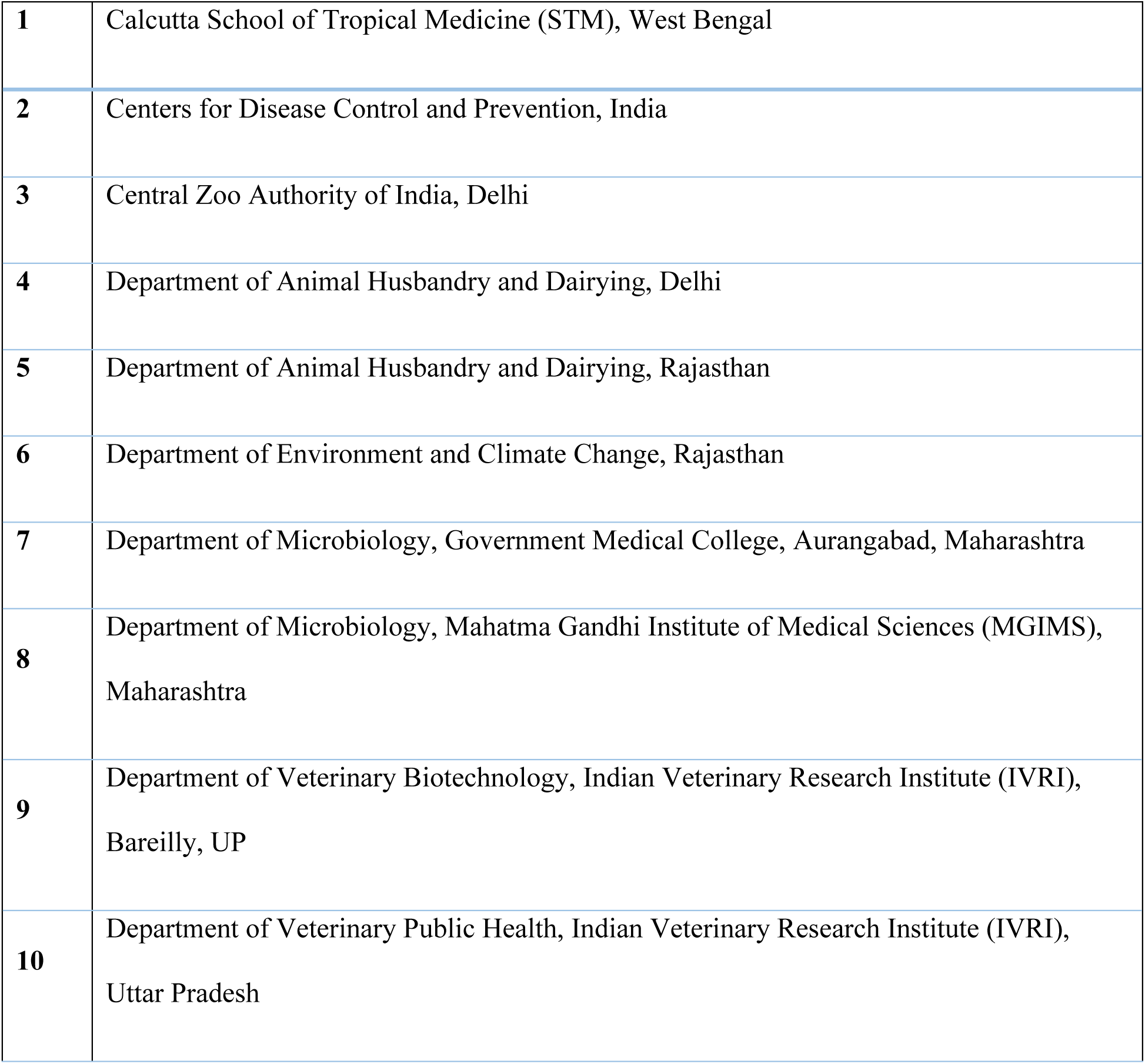
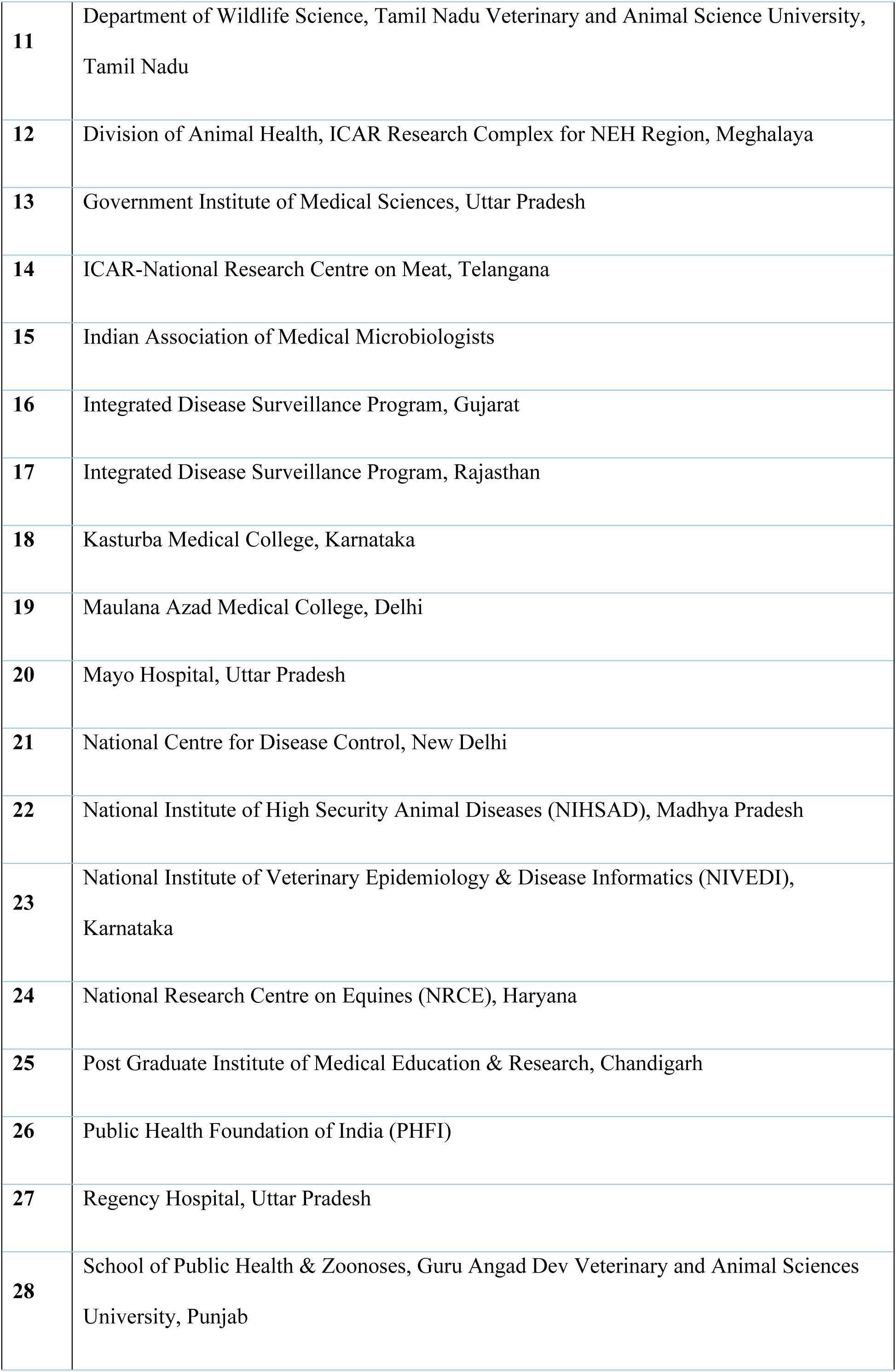
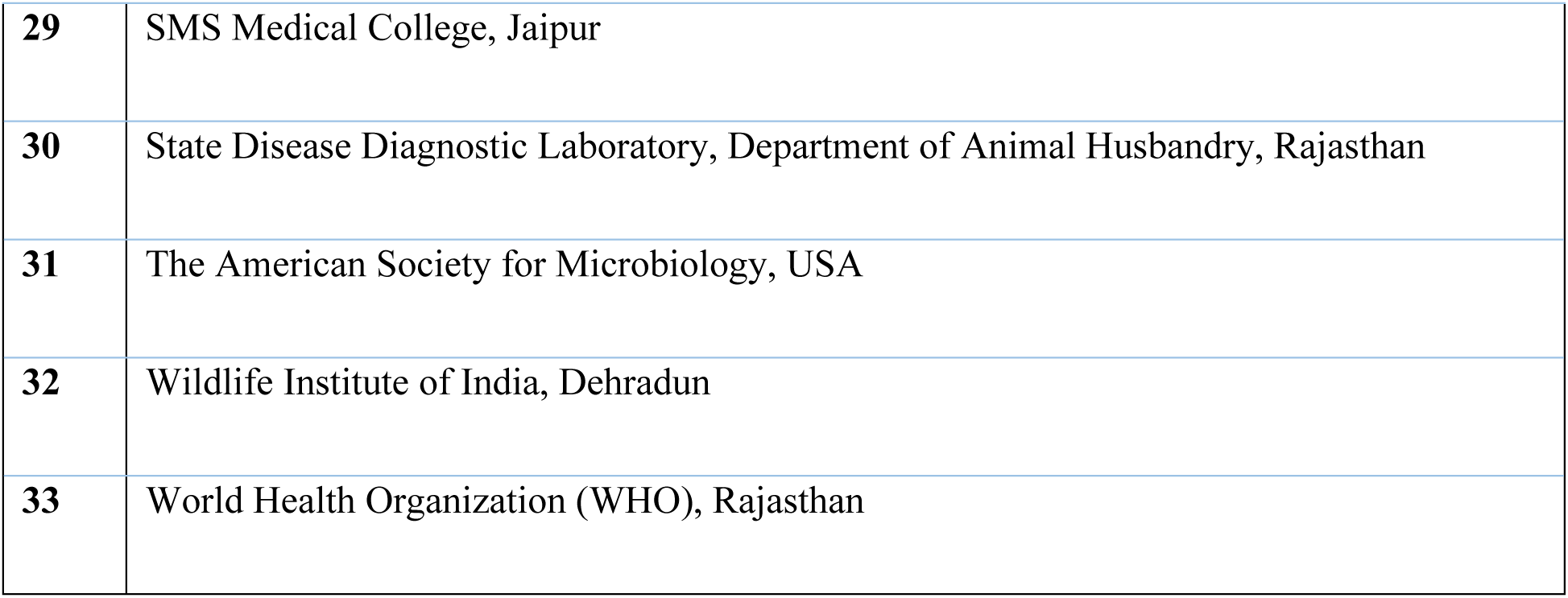
List of participating institutes and organizations.

**Table 2.** List of Zoonotic Diseases Considered for Prioritization.

|  |  |
| --- | --- |
| 1. Rabies | 21. Histoplasmosis |
| 2. Anthrax | 22. Zoonotic tuberculosis |
| 3. Brucellosis | 23. Salmonellosis |
| 4. Plague | 24. Melioidosis |
| 5. Leptospirosis | 25. Glanders |
| 6. Scrub typhus | 26. Cysticercosis |
| 7. Zoonotic Influenza (Zoonotic Influenza A viruses) | 27. Schistosomiasis |
| 8. Kyasanur Forest Disease (KFD) | 28. West Nile Virus |
| 9. Crimean-Congo hemorrhagic fever (CCHF) | 29. Q-Fever |
| 10. Nipah virus infection | 30. Leishmaniasis |
| 11. Japanese Encephalitis | 31. Listeriosis |
| 12. Toxoplasmosis | 32. Trypanosomiasis |
| 13. Campylobacteriosis | 33. MERS-coronavirus (MERS-cov) |
| 14. Lyme Disease | 34. Severe acute respiratory syndrome (SARS) |
| 15. Echinococcosis | 35. Ebola virus disease |
| 16. Dengue | 36. Yellow Fever |
| 17. Bartonellosis (Cat Scratch) | 37. Novel Coronavirus |
| 18. Psittacosis | 38. Lassa fever |
| 19. Tularemia | 39. Shiga Toxin-Producing E. Coli |
| 20. Cryptococcosis | 40. Blastomycosis |

**Table 3.**
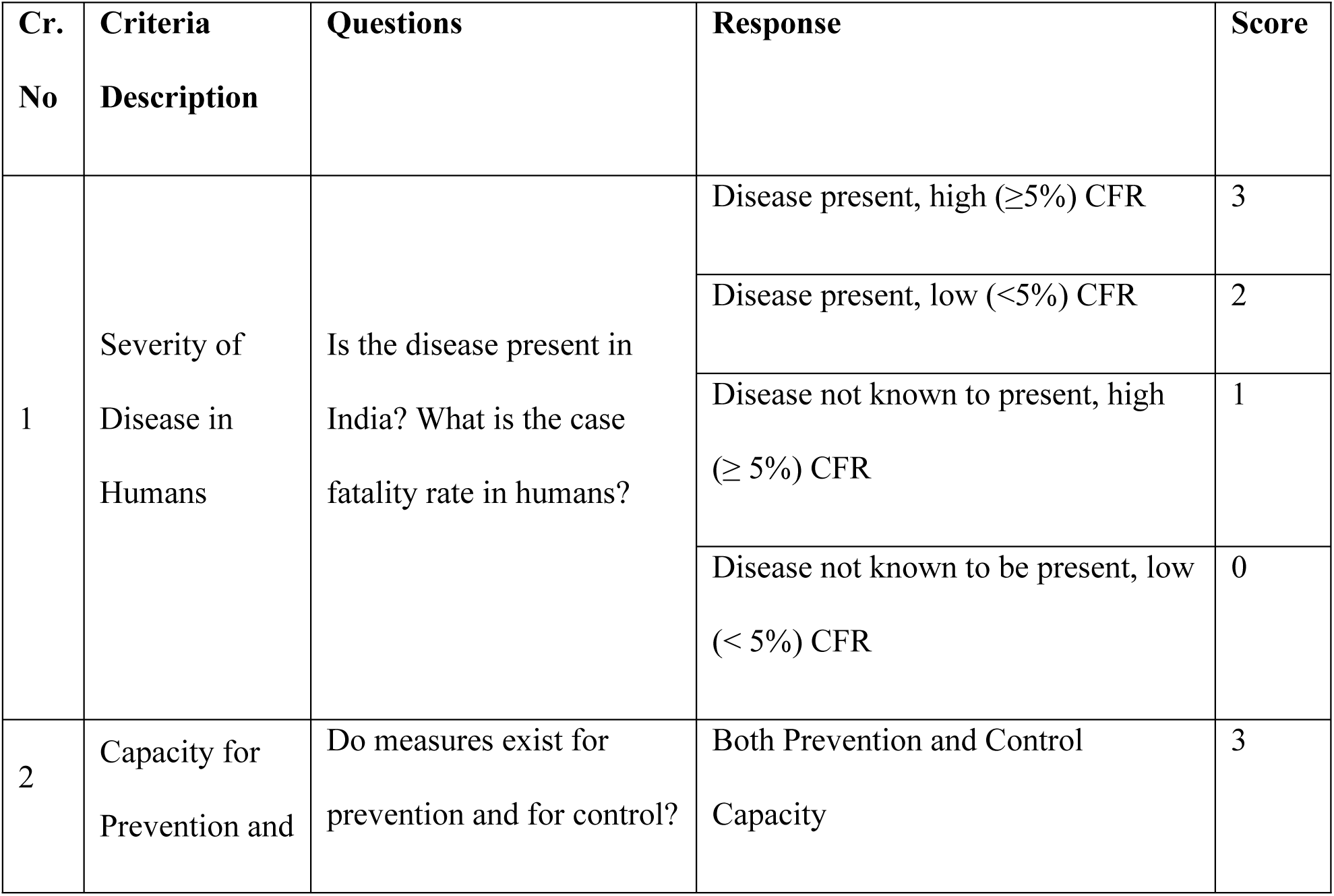
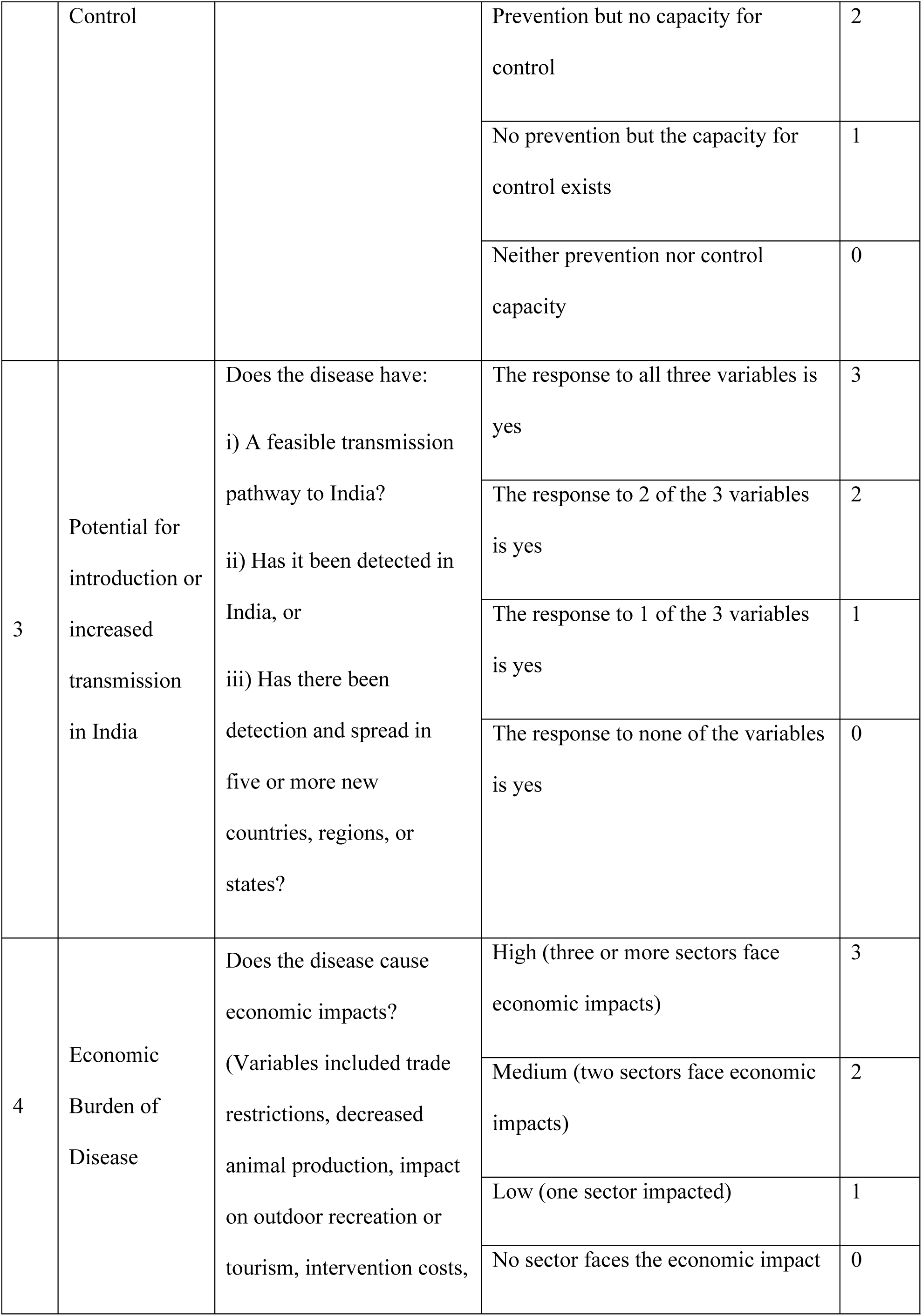
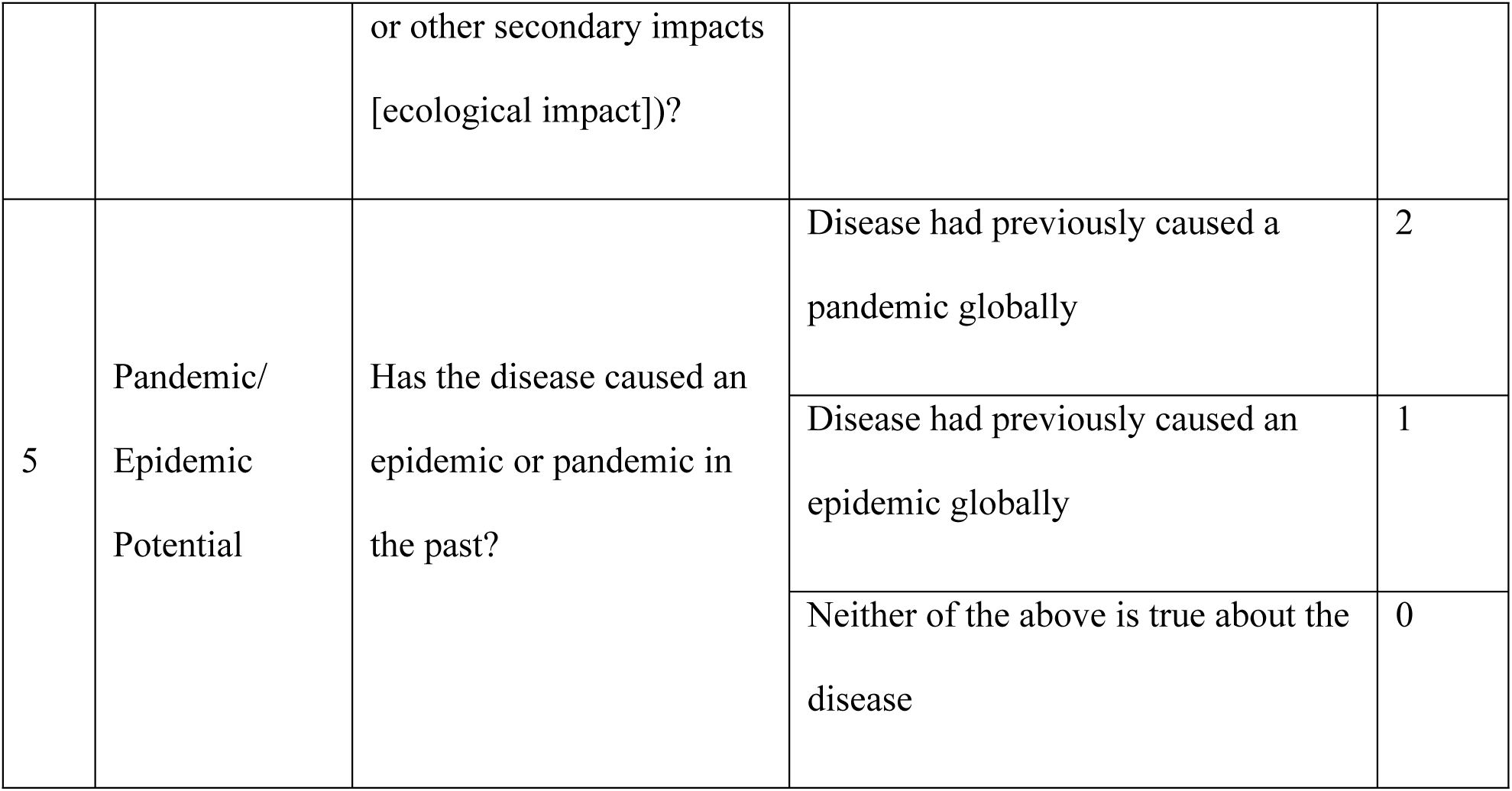
Criteria, Questions, Responses, and Scores Finalized for Ranking of Zoonotic Diseases.

### Step 4: Ranking of the criteria and weighted score calculation through the Analytic Hierarchy Process

On the first day of the workshop, the participants were familiarised with the Analytic Hierarchy Process (AHP), which was used to rank criteria and develop a weighted score for each criterion using the OHZDP Tool [12].

All the workshop participants were initially distributed in six sector-specific groups of 6-7 participants, a modification of CDC OHZDP methods. These groups ranked the five criteria finalized in step 2 to generate six sets of criteria rankings Table 4. The OHZDP Tool was then used to compute the average ranking from the six groups’ inputs and generate the final rankings and criteria weights.

**Table 4.** Group ranking of criteria for prioritizing zoonotic diseases using the Analytic Hierarchy Process.

| **Criteria** | **Group**<br>**1** | **Group**<br>**2** | **Group**<br>**3** | **Group**<br>**4** | **Group**<br>**5** | **Group**<br>**6** | **Overall**<br>**Ranking** |
| --- | --- | --- | --- | --- | --- | --- | --- |
| Severity of Disease in Humans | 0.18 (2) | 0.35 (1) | 0.46 (1) | 0.28 (2) | 0.35 (2) | 0.05 (5) | 0.45 (1) |
| Capacity for Prevention and Control in humans and animal health sector | 0.56 (1) | 0.35 (2) | 0.06 (5) | 0.20 (3) | 0.39 (1) | 0.49 (1) | 0.22 (2) |
| Potential of Transmission | 0.15 (3) | 0.09 (4) | 0.12 (3) | 0.28 (1) | 0.05 (4) | 0.16 (3) | 0.19 (3) |
| Economic Burden of Disease | 0.05 (5) | 0.10 (3) | 0.24 (2) | 0.07 (5) | 0.15 (3) | 0.08 (4) | 0.18 (4) |
| Pandemic or Epidemic Potential | 0.06 (4) | 0.09 (5) | 0.12 (3) | 0.18 (4) | 0.05 (5) | 0.22 (2) | 0.18 (5) |
| Consistency Ratio | 0.06 | 0.05 | 0.05 | 0.08 | 0.09 | 0.09 | N/A |

### Step 5: Scoring and final ranking of zoonotic weighted criteria

The voters, advisors, and facilitators were then reorganized into six heterogenous groups of representative stakeholders. Each group included a moderator from the US-CDC (India) or the American Society for Microbiology (ASM). Each of these groups were provided a list of 40 zoonotic diseases and a questionnaire to facilitate the scoring of each categorical question. They were also provided with references for each criterion identified from the literature review. Question scores were then entered into the OHZDP Tool to calculate final scores for each zoonotic disease, normalize these scores, and generate the final ranking for each of the 40 listed diseases Table 5. The OHZDP Process incorporated decision tree analysis to calculate the final weighted score by multiplying the weights of each criterion with the score assigned to each question. A consistency ratio of 0.1 or less was considered satisfactory [28]. We used an Excel-based program for AHP to rank the criteria and calculate the consistency ratio by performing pairwise comparisons of the criteria. For diseases for which participants felt that available data were too limited for scoring to be performed, ranks were given using simple decision rules based on heuristic knowledge of the existing capacity of healthcare systems, vaccination, and infrastructure in India. Ranks given using simple decision rules were confirmed by expert consensus within the groups. While this qualitative method had certain limitations in terms of its subjectivity, this was partially overcome by averaging the ranks from six groups and by utilizing the best available quantitative and qualitative data for expert consensus ranking.

**Table 5.** Normalized Scores for the forty Zoonotic Diseases Ranked based on OHZDP.

| S.No. | Name of Disease | Normalized Score |
| --- | --- | --- |
| 1 | Zoonotic Influenza (Zoonotic Influenza A viruses) | 1 |
| 2 | Anthrax | 0.9318421 |
| 3 | Japanese Encephalitis | 0.9146899 |
| 4 | Leptospirosis | 0.8946276 |
| 5 | Brucellosis | 0.8908413 |
| 6 | Dengue | 0.873689 |
| 7 | Rabies | 0.8636843 |
| 8 | Scrub typhus | 0.8460808 |
| 9 | Plague | 0.8447246 |
| 10 | Crimean-Congo hemorrhagic fever (CCHF) | 0.8114685 |
| 11 | Zoonotic tuberculosis | 0.793719 |
| 12 | Nipah virus infection | 0.793719 |
| 13 | Kyasanur Forest Disease (KFD) | 0.7850176 |
| 14 | Listeriosis | 0.7765668 |
| 15 | Salmonellosis | 0.7704677 |
| 16 | Echinococcosis | 0.7677194 |
| 17 | Campylobacteriosis | 0.7533154 |
| **18** | Q-Fever | 0.7355659 |
| **19** | Leishmaniasis | 0.7191725 |
| **20** | Glanders | 0.7084089 |
| **21** | West Nile Virus | 0.7079577 |
| **22** | Lyme Disease | 0.6991103 |
| **23** | Yellow Fever | 0.689026 |
| **24** | Shiga Toxin-Producing E. coli | 0.6847063 |
| **25** | Cysticercosis | 0.6505634 |
| **26** | Schistosomiasis | 0.6505634 |
| **27** | Toxoplasmosis | 0.6505634 |
| **28** | Trypanosomiasis | 0.6453343 |
| **29** | Melioidosis | 0.6397998 |
| **30** | Psittacosis | 0.6397998 |
| **31** | Cryptococcosis | 0.6397998 |
| **32** | Histoplasmosis | 0.6397998 |
| **33** | Blastomycosis | 0.6397998 |
| **34** | Bartonellosis (Cat Scratch) | 0.6305012 |
| **35** | MERS-coronavirus (MERS-CoV) | 0.5285279 |
| **36** | Severe acute respiratory syndrome (SARS) | 0.5285279 |
| **37** | Ebola virus disease | 0.5285279 |
| 38 | Novel Coronavirus | 0.5285279 |
| 39 | Tularemia | 0.484171 |
| 40 | Lassa fever | 0.3865318 |

Sensitivity analysis was conducted to test the validity of the prioritization results based on the impact of input variables (weighted criteria and scoring of individual zoonotic diseases). In the first stage of sensitivity analysis the five criteria were given an equal weight of 1 each to obtain normalized scores. In the next stage, scores for each of the zoonotic diseases were obtained by sequentially removing each of the five criteria. Pearson’s correlation was used to assess the relationship between normalized scores and the adjusted scores used to assess the impact of criteria weightage and their contribution to disease prioritization ranks. Pearson’s correlation coefficient was considered significant at p<0.05. The analysis was done in STATA software version 15.1.

### Step 6: Discussion on the way forward

The workshop concluded with a discussion on a road map for the prevention and control of identified priority zoonotic diseases as well as overall areas for improvement, including the development of a national one health action plan and robust linkages across all sectors.

### Deviations from the CDC OHZDP process and expected impact

This workshop followed standard CDC OHZDP methodology by using a list of zoonotic diseases developed in discussion with multiple stakeholders, utilizing five criteria to evaluate the diseases, using Excel-based tools for AHP to rank the criteria and Decision Tree analysis to derive weighted scores for each ranked criterion. Finally, weighted scores for each question were summed up to give the total score for each pathogen which were then normalized against the highest score to obtain the final ranked list. In addition, three groups of experts were created to act as advisors, facilitators and voting members. Some modifications were necessary, however, and they are described below.

#### Deviation 1

The criteria were decided before the workshop.

##### Deviation description and rationale

Development of the criteria, their responses, and the scores associated with each response were developed 2 days before the workshop. This was done through a videoconference during which the advisors, facilitators, and most of the voting members debated and ultimately achieved consensus on the criteria to be included, the responses, and the scores for each response. The discussion was led by NCDC, and prioritization documents of other countries were referenced to develop the final list. This was done to ensure the unhindered progress of the workshop without jeopardizing the timeline.

##### Impact on the process

This deviation limited the inclusion of all stakeholder perspectives in the development of criteria, which could have impacted criteria selection. This limitation was partially mitigated by presenting the list of criteria to the participants during the workshops for discussion and further suggestions. As there were no suggestions we proceeded to ranking. This modification allowed more time to be allotted for ranking and other steps.

#### Deviation 2

The voting members were divided into six sector-specific groups for criteria ranking.

##### Deviation description and rationale

There were 33 voting members available for ranking the criteria. While CDC recommends ranking by individual voters, we divided the voter cohort into 6 sector-specific groups with each group generating a rank for the criteria after discussion. These ranks were collated and used for AHP as outlined in Table 4. This was done as the number of voters exceeded the standard CDC OHZDP methodology. It was expected that sector-specific grouping would:

- Promote a knowledge-based discussion on each criterion before ranking
- Minimize score variation compared to individual scoring by 33 members
- Reduce the amount of time required for voting

The goal was to achieve a consistency ratio (CR) < 0.1 on AHP.

##### Impact on the process

A CR of < 0.1 was achieved for the ranking done by each group. As the values were much lower than the cutoff it can be concluded that the modified method had no negative impact on the process.

## Ethics Approval

This workshop did not require ethical approval or informed consent since it did not entail research on human subjects and any data on the workshop participants included in the final report was anonymized.

## Results

Out of the forty zoonotic diseases evaluated Table 2, the top ten ranked diseases were finalized Table 5. The following selected criteria, in the order of the value of their respective weights, were used to rank zoonotic diseases:

1. Severity of illness in humans – 0.45
2. Capacity for prevention and control in the human and animal sector – 0.22
3. Potential for introduction or increased transmission in India – 0.195
4. Economic burden of the diseases – 0.188
5. Pandemic potential – 0.187

The distribution of calculated weight and rank for each of the five criteria from each of the six groups are provided in Table 6.

**Table 6.** Definition and Final Weightage of the Criteria Identified for Zoonotic Disease Prioritization During the National One Health Workshop, February 2020.

| Criteria | Question asked | Response | Score |
| --- | --- | --- | --- |
| Severity of Disease in | Is the disease present in India? What is the case | Disease present, high CFR ($\geq 5\%$) | 3 |
| Humans<br>Weight: 0.45 | fatality rate in humans? | Disease present, low CFR (<5%) | 2 |
| | | Disease not known to present, high CFR ($\geq 5\%$) | 1 |
|  |  | Disease not known to be present, low CFR (< 5%) | 0 |
| Capacity for prevention and control<br>Weight: 0.22 | Do measures exist for prevention and for control? | Both Prevention and Control Capacity | 3 |
|  |  | Prevention but no capacity for control | 2 |
|  |  | No prevention but capacity for control exists = 1 | 1 |
|  |  | Neither prevention nor control capacity = 0 | 0 |
| Potential for introduction or increased transmission in India<br>Weight: 0.195 | Does the disease have:<br>1) A feasible transmission pathway to India?<br>2) Has it been detected in India? 3) Has there been detection and spread in five or more new countries, regions, or states? | The response to all three variables is yes | 3 |
|  |  | The response to 2 of the 3 variables is yes | 2 |
|  |  | The response to 1 of the 3 variables is yes | 1 |
|  |  | The response to none of the variables is yes | 0 |
| Economic Burden of Disease<br>Weight: 0.188 | Does the disease cause economic impacts? (Variables included trade restrictions, decreased animal production, impact on outdoor recreation or tourism, intervention costs, or other secondary impacts [ecological impact])? | High (three or more sectors face economic impacts) | 3 |
|  |  | Medium (two sectors face economic impacts) | 2 |
|  |  | Low (one sector impacted) | 1 |
|  |  | No sector faces the economic impact | 0 |
| Pandemic/Epidemic Potential<br>Weight: 0.187 | Has the disease caused an epidemic or pandemic in the past? | Disease had previously caused a pandemic globally | 2 |
|  |  | Disease had previously caused an epidemic globally | 1 |
|  |  | Neither of the above is true about the disease | 0 |

There was a strong positive correlation between normalized scores and adjusted scores computed by the OHZDP Tool by comparing weighted and unweighted criteria (r=0.95, p<0.05). There was a strong positive correlation when excluding each criterion from the model (r= 0.84-0.96, p < 0.05) (Fig 3). Sensitivity analysis demonstrated minimal changes in the disease ranking for the first 10 diseases in the final ranked list. Changes included 1) a drop in Brucellosis ranking from fifth to tenth, a drop in Rabies ranking from seventh to fourteenth, and a rise in plague ranking from ninth rank to second when the criterion “capacity for prevention and control in human and animal sector” was excluded; and 2) a drop in zoonotic influenza (zoonotic influenza A viruses) ranking from first to second when the criterion “pandemic and endemic potential” was excluded.

**Fig 3.**
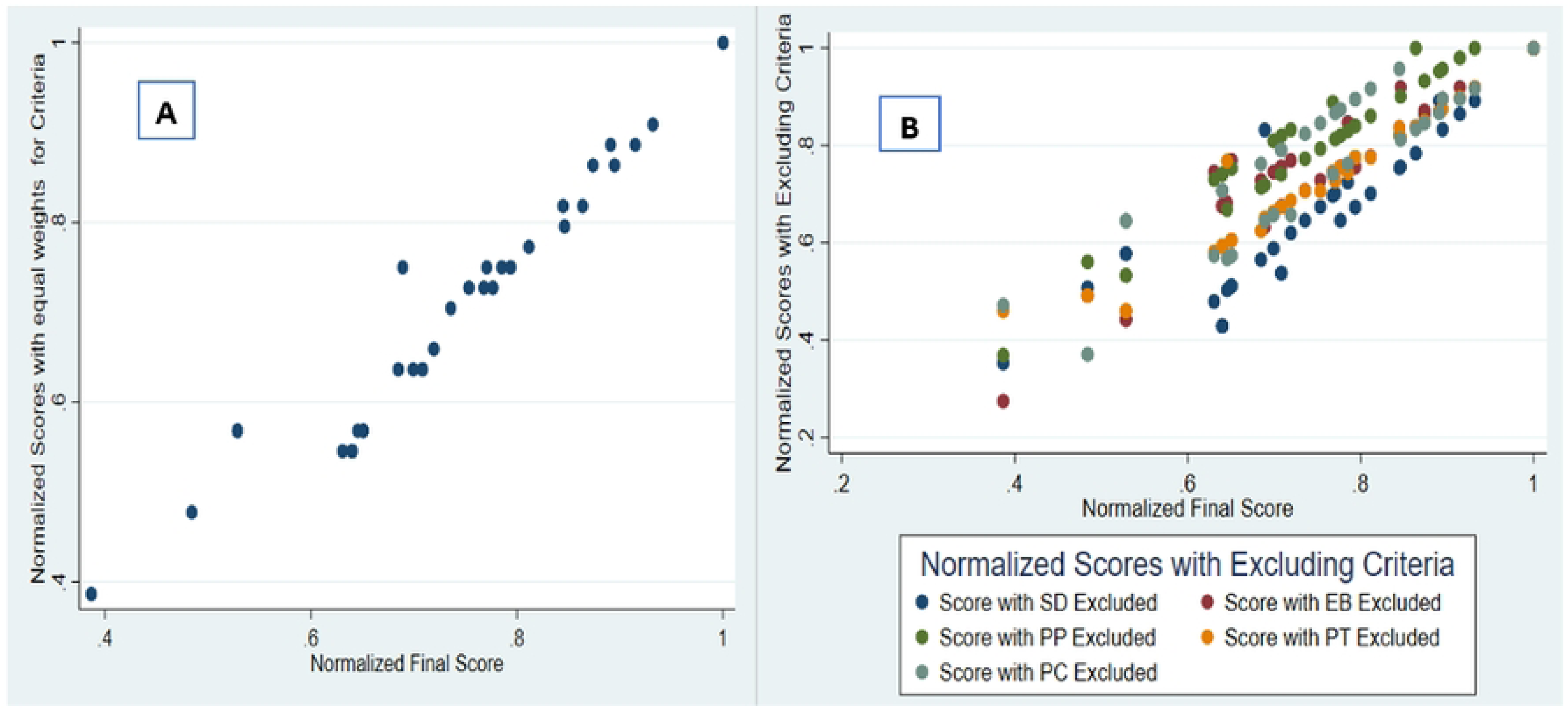
A Comparison of Normalized Scores Obtained from the Weighted Criteria and (A) Equal Weights (B) Excluding Each of the Five Criteria.

After the completion of the prioritization process, NCDC facilitated a discussion on a future roadmap for One Health in India. There was broad consensus on establishing clear communication channels between sectors and stakeholders, developing a joint reporting format, establishing a linked laboratory network across India at the state, district, and subdistrict levels, conducting joint surveillance and risk assessment, and institutionalization and implementation of practical One Health coordination mechanisms through the identification of clear roles and responsibilities for different ministries and levels of Government.

## Discussion

Through conduction of this prioritization workshop, India has demonstrated its commitment to One Health through inter-ministerial collaboration and a multi-disciplinary, multi-sectoral approach to prioritization demonstrating “functionality with bridges and fluidity” [29] in action. In a national symposium on One Health in India organized in 2021 [30], the Union

Minister of Health and Family Welfare announced a collaborative effort through the Indian Council of Medical Research (ICMR) and the Indian Council of Agricultural Research (ICAR) to develop a National Institute of One Health (NIO), which will support a high-level steering committee for Eco-Health Initiatives in India [30]. Both ICAR and ICMR were represented in this prioritization workshop and will be able to carry the discussions and agreed upon roadmap into the NIO and steering committee deliberations.

This workshop was the first time that stakeholders involved in the prevention and control of zoonotic diseases in humans, animals, wildlife and the environment at the national level in India used a prioritization process to collaboratively finalize a list of ten priority zoonotic diseases to chart out a future action plan. Previous efforts at the local level [25] identified Rabies, Brucellosis, avian influenza (H5N1), influenza A (H1N1), and Crimean-Congo haemorrhagic fever as the highest priority. That list closely resembles the results of this national-level workshop, with the exception of avian influenza which was not included in this workshop.

Prevention and control capacity and severity of disease in humans emerged as the two most important ranking criteria during this workshop. This was evident both in their respective weights and impact on ranking, with diseases like Zoonotic Influenza, Anthrax, and Japanese Encephalitis with existing prevention and control capacity receiving the highest scores and diseases like Lassa Fever and Tularemia, with little to no prevention and control capacity receiving the lowest scores.

This prioritization will be instrumental in furthering One Health in India, one example being the utilization of this prioritized list by the One Health Consortium (OHC). The OHC consists of medical centres, veterinary institutions, central government institutes, and eight disease investigation centres in the northeast states of India. The consortium is envisaged to initiate cross-cutting collaborations between animal, human, and wildlife health professionals. The objectives of this consortium are to establish a network of laboratories at centralized and field levels, estimate the prevalence and burden of selected priority diseases, detect pathogens by serological (antigen) or molecular tests, especially in clinical cases, and model data for disease forecasting as well as risk assessment. This consortium along with the National Institute of One Health (NIO) will work with the World Health Organization, several other health organizations, and national governments towards the successful application of One Health principles [31]. A second example of current application of prioritization results for further One Health action is The National One Health Program for Prevention and Control of Zoonosis’ (NOHPPPCZ’s) use of workshop results to enable rapid, timely detection and reporting of zoonotic diseases through a network of sentinel surveillance sites for zoonotic diseases and a comprehensive One Heath action plan for the detection, prevention, and control of these diseases. A data sharing mechanism leveraging the existing Integrated Health Information Platform will be invaluable in achieving enhanced surveillance.

Limitations of this prioritization process include underreporting of zoonotic diseases in formal reporting mechanisms and associated selection bias. These limitations were partially overcome by an extensive literature review conducted before the workshop, and with the use of a mixed-methods approach to fill the quantitative gaps with qualitative inputs through expert consensus. Limitations imposed by deviations in OHZDP methodology are addressed in the Methods section.

### Conclusions

Zoonotic influenza (zoonotic influenza A viruses) was the highest ranked zoonosis in this workshop. Overcoming the programmatic silos that currently exist within different government agencies and institutions will be a major challenge in operationalizing One Health in India, however, this workshop demonstrated that such coordination is possible. This prioritization conducted at the national level has the potential to catalyse such efforts at the state and local levels across India by fostering the communication, collaboration, cooperation, and coordination necessary to make meaningful progress. Given the diversity in geography and prevalence of communicable diseases in different parts of India, states may be interested in taking up similar exercises to further streamline their needs and resource utilization. State action plans developed under the guidance of the National One Health Program for Prevention and Control of Zoonoses (NOHPPCZ) will help the states achieve the one health goals outlined by the program.

## Data Availability

All relevant data are within the manuscript and its Supporting Information files.

## Acknowledgement

The authors would like to thank all the stakeholders and experts from the Government of India, Directorate of Medical and Health Services, Rajasthan, and National and State level institutions for participating in the workshop, as well as the US-CDC India country office for organizing and coordinating the session.

## Disclaimer

The opinions expressed by authors contributing to this article do not necessarily reflect the opinions of the Ministry of Health and Family Welfare, Ministry of Fishery, Department of Animal Husbandry and Dairying, Ministry of Agriculture and Farmer’s Welfare, India, U.S. Centers for Disease Control and Prevention, or any other institutions with which the authors are affiliated.

